# Validity of self-screening with the KOJI AWARENES™ test for range of motion and strength in healthy subjects

**DOI:** 10.1101/2024.04.24.24306281

**Authors:** Kenji Hirohata, Hidetaka Furuya, Sho Mitomo, Yuki Osaka, Koji Murofushi, Kazuyoshi Yagishita

## Abstract

**Objective:** This study aimed to establish the validity of the KOJI AWARENESSTM sub-components by determining whether there is a connection between the sub-component scores and joint range of motion, muscle strength, and balance.

**Methods:** Fifty healthy adults (17 females and 33 males) participated in the study, completing both the KOJI AWARENESS^TM^ and measurements of joint range of motion, muscle strength, and balance. The range of motion of the upper and lower extremities and trunk was measured using either a goniometer or an inclinometer. A handheld dynamometer was used to measure muscle strength. Balance ability was assessed using a modified balance error scoring system. Using the Mann–Whitney U test or Jonckheere–Terpstra test, we compared the KOJI AWARENESS™ score and the corresponding body segments, with a significance level of P ≤ 0.05.

**Results:** Our results indicated that there were associations between external references and many items, but no associations were found for flexion, extension, and rotation of the "neck mobility," extension and external rotation of the "hip mobility," and strength of the "mid-section stability strength" in the KOJI AWARENESS^TM^.

**Conclusion:** Overall, the KOJI AWARENESS^TM^ sub-component scores showed good validity, with the exception of the items related to neck and hip flexibility and trunk muscle strength. Future analyses should include a wider range of age groups, such as middle-aged and elderly individuals.

## Introduction

Self-checking of physical function and health status can help prevent disease and injury [1–5]. We developed the KOJI AWARENESS^TM^, a self-screening test to assess musculoskeletal functions, including flexibility and muscle strength. The Koji Awareness screening test is fundamentally designed to screen motor function in a holistic and composite manner [6]. Our previous research confirmed that the KOJI AWARENESS^TM^ is a valid test that correlates strongly and positively with scores on the Functional Screening Test (FMS), a well-known screening test for motor function [7]. The FMS can be used to predict injury and has been demonstrated to have adequate reliability and validity in systematic reviews [8–10]. Thus, the KOJI AWARENESS^TM^ is indicated to adequately assess an individual’s motor function to some extent.

The KOJI AWARENESS^TM^ is a self-screening test comprising 11 components, which include neck mobility, shoulder mobility, shoulder blade mobility, thoracic spine mobility, upper extremity stability/strength, hip mobility, hip/spine mobility, upper extremity mobility and stability, midsection stability strength, lower extremity strength, and ankle mobility. Each component is rated on a 2–4 Likert scale. For all components, a higher score indicates a better state of physical function; however, it is unclear which specific physical function (joint range of motion (ROM), muscle strength, etc.) is associated with each component score. The validity of the KOJI AWARENESS^TM^ total score regarding the FMS score has been demonstrated; however, the validity of the sub-components remains unclear [7].

Therefore, this study aimed to clarify the validity of the KOJI AWARENESS^TM^ sub-components by confirming the relationship between their scores and joint ROM, muscle strength, and balance. We hypothesized that the KOJI AWARENESS™ sub-components would have some validity in relation to their corresponding ROM, muscle strength, and balance.

## Methods

### Subjects

Fifty healthy adults (17 females and 33 males) participated in this cross-sectional study. All participants were recruited between June and August 2021. Subjects were included if they met the following criteria: (1) healthy, with no limitations in daily life activities; (2) aged between 20 and 60 years; and (3) no severe injuries in the last 3 months. Subjects were excluded if they met any of the following conditions: (1) severe psychiatric, neurological, or cardiovascular disease; (2) orthopedic disorder; (3) pregnancy; or (4) acute infectious disease. Prior to the measurement, all subjects provided written informed consent to participate in the study. The participants were instructed to stop when they experienced pain or discomfort during any part of the test. This study was approved by the Research Ethics Committee of Tokyo Medical and Dental University (research protocol identification number: M2021-029) and followed the Declaration of Helsinki Ethical Principles (52nd WMA General Assembly, Edinburgh, Scotland; October 2000) for medical research involving human subjects. All 50 participants completed the evaluations described below.

### Demographic characteristics

Participation in any type of exercise and/or sporting activity was recorded. Age, sex, height, and weight were recorded on the testing day. Body mass index (BMI) was calculated based on each participant’s height and weight.

### Movement screening tests: KOJI AWARENESS™

The KOJI AWARENESS^TM^ is composed of measures of ROM, muscle strength, and balance [7]. Further details on KOJI AWARENESS™ are provided in Appendices 1 and 2. The participants used a checklist to self-evaluate the function of each body part. There were 11 designated movements for self-evaluation, and each component had distinct scoring criteria, with a maximum total score of 50 points. Each component of KOJI AWARENESS^TM^ was divided to reflect the corresponding segments of the body so that the subjects could immediately locate the dysfunctional body region. The KOJI AWARENESS ™ method was explained to the participants until they understood it. Subsequently, they self-rated the motor function of each item according to the method presented in Appendices 1 and 2. Unilateral and asymmetrical tests were performed on both sides of the body. Up to three attempts were allowed, and the best score was retained. The participants completed the assessment within an average of 20 min. To improve reproducibility, all subjects completed the KOJI AWARENESS™ with guidance from the same athletic trainer (ATC) who was certified by the Board of Certification, Inc.

## External references

### Range of motion and flexibility

A universal goniometer was used to measure the ROM of each body segment, including the cervical spine (flexion, extension, lateral flexion, and rotation), shoulder joint (abduction), thoracic spine (rotation while sitting), and hip joints (flexion, extension, and rotation). An internal rotation behind-the-back angle test [11] was performed to assess glenohumeral internal rotation flexibility. With the subjects in a standing position, they were instructed to reach the highest point along the midline. The internal rotation behind-the-back angle was defined as the angle between the ulna and the line of gravity. To measure the internal rotation behind-the-back angle, we used a goniometer and measured in 5-degree units. Active spinal flexion and extension mobilities were measured while the subjects were standing. Subjects were asked to flex or extend their spines as far as possible. Measurements were performed in the active end-range position. Spinal flexion and extension mobilities were measured using an inclinometer. The difference in angle between the spinous process of the 1st thoracic and 1st sacral spine was recorded.

The straight leg raise (SLR) test was performed [12, 13]. With the subject in the supine position and the opposite leg attached to the table, compensation was minimized. The tester lifted the leg off the table while the knee was extended. The endpoint for straight leg raising was determined by one or more of three criteria: (1) the knee started to flex, (2) the tester perceived firm resistance, and (3) palpable onset of posterior pelvic rotation. At the endpoint, hip ROM was recorded using a goniometer.

The weight-bearing lunge test was also performed [14]. The subjects were positioned facing a wall, with the line connecting the second toe and heel of the test foot perpendicular to the wall. While maintaining this position, the subjects were instructed to perform a lunge in which the knee was flexed with the goal of making contact between the knee cap and wall.

### Muscle strength

A handheld dynamometer (Mobie MT-100B, Sakai Med, Tokyo, JAPAN) was used to measure isometric muscle strength, shoulder abduction, trunk flexion, and knee extension. To measure shoulder abduction muscle strength, we referred to and modified the method described by Kibler et al.[15]. The subjects were instructed to hold their arm in the test position (90° abduction in the scapular plane and shoulder external rotation) and to provide a one-repetition maximum voluntary isometric contraction against resistance in that position. Isometric trunk flexion muscle strength was measured in a sitting position with the knee flexed at 90° and the back attached to the wall. To maintain the posterior pelvic tilt, measurements were taken with a towel set between the buttocks and the wall, with the back in contact with the wall. A dynamometer was placed on the sternum. The subjects were instructed to place their hands in front of the chest and to gradually increase isometric trunk flexion strength for 3 s.

To measure knee extension strength, we modified the method described by Hansen et al.[16]. The subjects were seated with their knees flexed at 90° and their arms crossed in front of their chest. A strap was used to fix the thighs to the seated surface. Another strap was attached to the leg of the table to stabilize the handheld dynamometer during the measurement. The subjects were instructed to gradually increase their isometric knee extension strength for 3 s. The measurements were performed three times, and the average value was used for the analysis.

### Balance

We used the modified Balance Error Scoring System (mBESS) to evaluate postural stability [17]. The mBESS protocol comprises four conditions: feet together, single-leg stance, and tandem stance on firm and foam surfaces. The subjects were instructed to close their eyes and place their hands on their hips throughout each trial. Upon loss of balance, participants were instructed to return to their position as quickly as possible. The testers counted the number of errors during the 20-s trial. An error was defined as opening the eyes, stepping, lifting hands off the hips, lifting the forefoot or heel, abducting the hip by > 30°, stumbling or falling out of position, or failing to return to the test position in <5 s [18]. The maximum total error for each 20-s condition was 10, with a maximum error score given to subjects who could not maintain a position for a minimum of 5 s for each stance.

### Statistical analysis

The normality of the distribution of each variable was confirmed using histograms and the Shapiro–Wilk test. The mean ± standard deviation was used to summarize normally distributed data, and the median (interquartile range) was used for data that were not normally distributed. External references that matched each sub-component of KOJI AWARENESS^TM^ were selected and analyzed.

The Mann–Whitney U test was used to compare the differences between each component of the KOJI AWARENESS™ score and the corresponding body segment (P ≤ 0.05). The Jonckheere–Terpstra test was used to compare the KOJI AWARENESS™ score and more than two corresponding body segments (P ≤ 0.05). SPSS software (version 21.0; IBM Corp., Armonk, NY, USA) was used for all data analyses.

## Results

Fifty subjects (men = 34, age = 27 ± 5.4 years, height = 172.5 ± 5.7 cm, weight = 70.4 ± 12.0 kg, BMI = 23.6 ± 3.7 kg/m^2^; women = 16, age = 26 ± 4.7 years, height = 161.9 ± 4.6 cm, weight = 53.8 ± 5.4 kg, BMI = 20.6 ± 2.5kg/m^2^) participated in this study. The average KOJI AWARENESS™ score was 41.6 ± 5.8. We found associations between external references and many items; however, no associations with external references were found regarding flexion, extension, and rotation of the “neck mobility,” extension and external rotation of the “hip mobility,” and strength of the “mid-section stability Strength” in the KOJI AWARENESS^TM^(Table 1).

**Table 1.** KOJI AWARENESS^TM^ scores.

| Component of KOJI AWARENESS |  | External references | P value |
| --- | --- | --- | --- |
| **1. Neck mobility** |  |  |  |
|  | forward bending* | cervical flexion ROM | 0.131 |
|  | backward bending* | cervical extension ROM | 0.743 |
|  | side bending* | cervical lateral flexion ROM | 0.001 |
|  | rotation* | cervical rotation ROM | 0.395 |
| **2. Shoulder mobility\*** |  | Internal Rotation Behind-the-Back Angle | <0.001 |
| **3. Shoulder blade mobility\*** |  | shoulder abduction ROM | 0.002 |
| **4. Thoracic spine mobility§** |  | thoracic rotation ROM | 0.01 |
| **5. Upper extremity stability and strength§** |  | shoulder abduction muscle s strength | 0.008 |
|  |  | isometric trunk flexion muscles strength | 0.029 |
| **6. Hip mobility** |  |  |  |
|  | flexion-internal rotation* | hip flexion ROM | 0.025 |
|  |  | hip internal rotation ROM | <0.001 |
|  | flexion-external rotation* | hip flexion ROM | 0.022 |
|  |  | hip external rotation ROM | 0.045 |
|  | extension-internal rotation* | hp extension ROM | 0.001 |
|  |  | hip internal rotation ROM | 0.001 |
|  | extension-external rotation* | hip extension ROM | 0.971 |
|  |  | hip external rotation ROM | 0.72 |
| **7. Hip and spine mobility** |  |  |  |
|  | forward bending§ | hip flexion ROM | 0.01 |
|  |  | straight leg raise test | <0.001 |
|  |  | spinal flexion mobility | 0.61 |
|  | backward bending§ | shoulder abductor ROM | 0.051 |
|  |  | hip extension ROM | 0.346 |
|  |  | spinal extension mobility | 0.013 |
| **8. Upper and lower extremity mobility and stability§** |  | hip flexion ROM | 0.001 |
|  |  | hip extension ROM | 0.001 |
|  |  | mBESS score | 0.485 |
| **9. Mid-section stability strength§** |  | lumber flexion mobility | 0.581 |
|  |  | isometric trunk flexion muscles strength | 0.256 |
| **10. Lower extremity strength§** |  | mBESS score | <0.001 |
|  |  | isometric knee extension strength | 0.001 |
|  |  | WBLT | 0.052 |
| **11. Ankle mobility\*** |  | WBLT | <0.001 |
| *The Mann-Whitney U test |  |  |  |
| §The Jonckheere-Terpstra test |  |  |  |

## Discussion

This study aimed to clarify the validity of the KOJI AWARENESS^TM^ score sub-components by confirming the relationship between the scores of the sub-components and the joint range of motion and muscle strength that could be associated with each component. The results showed that the KOJI AWARENESS^TM^ sub-component scores generally had good validity, except for items related to neck and hip flexibility and trunk muscle strength.

### Neck mobility

Among the items assessing neck flexibility, the side-bending scores of the KOJI AWARENESS^TM^ and goniometric measurements showed an association. In contrast, no association with the goniometric measurements was found for forward bending, backward bending, or neck rotation. Goniometric active cervical range measurements, applied as external references, have been reported to have adequate measurement reproducibility [19]. The median (interquartile range) ROM values collected in the present study were 54.0° (22.3), 69.0° (18.1), and 66.3° (12.1) for anterior flexion, backward flexion, and rotation of the neck, respectively. There are some variations among reports regarding normative data on cervical motion [20]. Our results were similar to those of previous studies [21, 22] that measured cervical ROM using goniometers and compasses, as in the current study. No significant differences were found in the data summarized by dividing the groups based on the KOJI AWARENESS^TM^ neck flexibility scores. This study included healthy participants within a narrow age range, which may have biased the results. In the future, it will be necessary to collect and validate data from the elderly and those with cervical symptoms and limited ROM.

### Shoulder mobility/shoulder blade mobility

The KOJI AWARENESS^TM^ shoulder mobility score is associated with internal rotation behind-the-back angle. Similarly, scoring of “shoulder blade mobility” was also associated with maximum shoulder joint abduction angle. In scoring “shoulder mobility,” points are determined by the ability to touch the opposite shoulder blade with the hand behind the back. In scoring “shoulder blade mobility,” points were determined by whether the maximum abduction position could be maintained. Among young healthy adult men and women in this study, the median difference in shoulder internal rotation ROM between those with a “shoulder mobility” score of 1 point and those with a score of zero points was approximately 40°. The median difference in shoulder abduction ROM between patients with “shoulder blade mobility” scores of 1 point and those with a score of zero points was approximately 7°. These scores are valid as assessments that reflect the degree of shoulder internal rotation and abduction ROM. However, this study did not include elderly patients or those with a history of shoulder disease. Shoulder joint ROM decreases with age [23]. The “shoulder mobility” and “shoulder blade mobility” scores of the KOJI AWARENESS^TM^ may indicate a floor effect when targeting individuals with shoulder ROM limitations due to aging or other reasons. In such cases, it may be necessary to modify the rating to ensure that it is responsive to those with shoulder ROM limitations, such as the addition of another grading level.

### Thoracic spine mobility

In this study, the median thoracic rotation angle, measured using a goniometer, was 51.8° (20.8). Summarized using the KOJI AWARENESS^TM^ scoring system, the median values were 38.3°, 49.0°, and 56.5° for 1, 2, and 3 points, respectively. Our evaluation of thoracic rotation angle measurements in the lumbar locked position has been proven to be sufficiently reliable [24]. Johnson et al. measured thoracic rotation angles in 46 healthy adults in the lumber locked position and reported a mean value of 40.8 ± 10.7° [24]. Using a similar method, Furness et al. reported a mean thoracic rotation angle of approximately 41° in 12 healthy male and female subjects [25]. Our measurement results were similar to those of previous studies and were considered valid. The KOJI AWARENESS^TM^ score for “thoracic spine mobility” was determined on a four-point scale from 0 to 3, with difficulty adjusted by varying the upper extremity position. As the score increased by 1 point, the thoracic rotational ROM increased by approximately 7°. The KOJI AWARENESS^TM^ scoring for “thoracic spine mobility” was statically found to reflect the actual thoracic rotation angle and is a valid ordinal scale to determine the thoracic flexibility of the subject.

### Upper extremity stability and strength

The KOJI AWARENESS^TM^ scoring for “upper extremity stability and strength” was based on four types of posture-holding ability. This score is associated with isometric shoulder abduction and trunk flexion muscle strength. In isometric shoulder abduction, the deltoid and serratus anterior muscles act as the primary muscles [26, 27]. Abdominal muscle activity is important for isometric trunk flexion [28, 29]. In the position used for KOJI AWARENESS^TM^ scoring, the activity of the serratus anterior muscles, which act on scalene protraction, and the abdominal muscle group, which stabilizes the trunk against gravity, is important [26]. In particular, the closer the trunk approaches the horizontal and the longer the lever arm, the greater the activity of the serratus anterior and abdominal muscle groups required to maintain posture [30]. The KOJI AWARENESS^TM^ score for “upper extremity stability and strength” reflects the function of the shoulder abductor and trunk flexor muscles to some extent. However, all subjects in this study scored >3 points, and none of the subjects scored 0–2 points. Future analyses should include data on targets that correspond to 0–2 points to confirm the validity of the level setting. The present study confirmed the sequence of 3 and 4 points in the KOJI AWARENESS^TM^ scoring for “upper extremity stability and strength”.

### Hip mobility

In the KOJI AWARENESS^TM^ scoring for “hip mobility,” hip flexibility was assessed using a two-plane combined motion. The patterns included flexion–internal rotation, flexion–external rotation, extension–internal rotation, and extension–external rotation. The patterns of flexion–internal rotation, flexion–external rotation, and extension–internal rotation were associated with goniometric ROM measurements; however, no association was observed with hip extension–external rotation. An investigation of 120 healthy adults, including both males and females, aged 22–60 years (mean = 39.1 years), showed that the mean ROM of external rotation of the hip joint in hip extension was 41.8 ± 10.2° [31]. The mean value in the current study was 53.1 ± 9.8°, which is higher than that reported in previous studies. The participants in this study were in their 20s, and the fact that they were younger than those in previous studies may have influenced the results. In addition, patients with strong anteversion of the femur and hip had significantly reduced ROM of external rotation during hip extension [32]. In the study population, those with a history of developmental dysplasia of the hip and those with hip symptoms were excluded. Therefore, patients with general hip flexibility were included.

Therefore, it is possible that there was less variation in the ROM data, and fewer points were deducted in the KOJI AWARENESS^TM^ scoring, resulting in no association. Future studies should be conducted on those with decreased hip extension and external rotation ROM, such as the elderly and/or those who present with hip symptoms.

### Hip and spine mobility: forward bending

The KOJI AWARENESS^TM^ score for “hip and spine mobility (forward bending)” was associated with goniometric hip flexion ROM and passive SLR angle. A similar assessment to the KOJI AWARENESS^TM^ score is the finger–floor distance (FFD), which measures the distance between the fingertips and the floor when the trunk is bent forward to the maximum extent possible while holding the standing knee joint extension position. A previous study [33] reported that FFD was more strongly associated with pelvic motion than with lumbar motion during forward bending. In other words, FFD is thought to reflect hip flexibility rather than spinal flexion. Another report indicated that pelvic motion during forward bending was reduced when the passive SLR angle was reduced [34]. The data obtained in our study support those of previous studies; KOJI AWARENESS^TM^ scoring for “hip and spine mobility” could be an alternative assessment method to FFD.

### Hip and spine mobility: backward bending

The KOJI AWARENESS^TM^ score for “hip and spine mobility (backward bending)” is associated with shoulder abduction ROM and spinal extension mobility. The KOJI AWARENESS^TM^ scoring involves upper extremity elevation; therefore, shoulder abduction ROM may be relevant. In this study, spinal extension mobility was measured as an external criterion using dual inclinometry of the first thoracic and first sacral spine angles in the maximum extension position. The median spinal extension mobility for the current study subjects was 29.8°, 40.0°, and 55.5° for the 1-, 2-, and 3-point KOJI AWARENESS^TM^ backward bending scores, respectively. Better-performing patients had larger spinal extension angles, with a difference of approximately 10-15° between each point. The KOJI AWARENESS^TM^ backward bending score was found to reflect spinal extension mobility.

### Upper and lower extremity mobility and stability

As the KOJI AWARENESS^TM^ scoring for “upper and lower extremity mobility and stability” requires holding the posture in the single-leg standing position, we analyzed the relationship between this score and hip extension/flexion ROM and mBESS. The score was associated with hip extension/flexion ROM; however, it was not associated with mBESS scores. The posture required by the test requires mobility of hip flexion on the raised side and hip extension on the supporting side. Therefore, this scoring may have been associated with hip extension/flexion ROM. Regarding balance, Iverson et al. found that BESS scores progressively increased with age in a study of adult men and women aged 20–69 years [35]. In our study, the participants were limited to those who were approximately 27 years old, which may have reduced the variability in the mBESS scores and may not have been associated with the KOJI AWARENESS^TM^ scoring for “upper and lower extremity mobility and stability.” Future analyses should target a wide range of age groups, including middle-aged and elderly individuals.

### Lower extremity strength

In the KOJI AWARENESS^TM^ score, “lower extremity strength” was assessed with single-leg standing from the chair seated or half-kneeling position and was scored on a five-point Likert scale of 0–4 points. The higher the score, the better the balance and the greater the knee extensor strength. Kishigami et al. reported that the ability of elderly Japanese individuals to stand up from a chair is related to the cross-sectional area of the quadriceps muscle [36]. Our results support the findings of this previous study. Single-leg standing tasks from a chair are used in a wide variety of fields, including sports science and geriatrics [36–39]. However, for some subjects, such as the elderly, the difficulty level may be too high. Therefore, the exercise task was set from half-kneeling at one or two points on the KOJI AWARENESS^TM^ scoring, which makes the scoring system more adaptable to subjects with relatively low physical function. In this study, the number of participants who scored 0–2 points was limited because of their relatively young age. Future analyses should include data from elderly individuals.

### Ankle mobility

The KOJI AWARENESS^TM^ score for “ankle mobility” was associated with the dorsiflexion angle on the weight-bearing lunge test (WBLT). The WBLT is one of the leading methods for evaluating ankle dorsiflexion flexibility in the weight-bearing position [40–42]. In the WBLT, the tibial forward tilt angle or toe–wall distance is commonly measured in the maximal dorsiflexion position [40, 42]. The criterion used in the KOJI AWARENESS^TM^ scoring was whether the toe–wall distance was greater than or equal to the subject’s knuckle. Langarika-Rocafort et al. reported that the tibial forward tilt angle and toe–wall distance during the WBLT are correlated [43]. The results of the current study support those of previous studies. Thus, ankle mobility scoring in KOJI AWARENESS^TM^ was found to be a valid method.

### Clinical implication

The KOJI AWARENESS^TM^ was designed to enable a self-check of physical functions, such as flexibility, muscle strength, and balance. Self-checking of health and physical condition leads to positive lifestyle changes. Self-monitoring of weight and food intake is a lifestyle therapy recommended for obese individuals [44, 45]. Feedback on sleep parameters using personal health monitors improves sleep outcomes [46]. Self-monitoring has been used to monitor changes in physical activity [47]. Thus, awareness of one’s own condition increases intrinsic motivation and causes behavioral changes. This may also be true for flexibility, muscle strength, and balance. The results of this study confirm that KOJI AWARENESS^TM^ sub-component scoring is, to some extent, valid for assessing physical function. Various researchers have reported that a decline in musculoskeletal functions, such as flexibility, muscle strength, and balance, is a risk factor for physical pain and orthopedic disorders [48–51]. If KOJI AWARENESS^TM^ becomes commonplace and the practice of self-assessing and improving the scores of musculoskeletal and other physical functions becomes widespread, it can be expected to reduce the risk of developing pain and orthopedic diseases.

### Limitations

This study had several limitations. First, there were several sub-components of KOJI AWARENESS^TM^ that could not be validated. Second, only healthy young adults were recruited; additional studies are needed to include older adults and those with orthopedic conditions for all sub-components. Finally, only Japanese patients were included in this study; however, different races have different bone morphologies and limb lengths relative to body height [52]. In the KOJI AWARENESS^TM^ scoring, several components are affected by upper and lower extremity lengths, and the effect of these physical differences may lead to different results when surveyed in other racial groups. Therefore, caution should be exercised when applying the results of this study in other countries.

## Data Availability

All relevant data are within the manuscript and its Supporting Information files.

## Acknowledgments

The authors thank all the participants who were included in this study. We thank Editage for editing and reviewing this manuscript for English language. This work was supported by MHLW Program (Grant Number 22JA1006).

## Supporting Information

**S1 Appendix. KOJI AWARENESS^TM^ movement test.**

**S2 Appendix. Scoring and criteria of KOJI AWARENESS^TM^**

